# Metabolic syndrome and hemorrhagic stroke among symptomatic CCMs in the Mexican Hispanic Population

**DOI:** 10.1101/2023.05.15.23289984

**Authors:** Alok K. Dwivedi, David Jang, Ofek Belkin, Justin Aickareth, Mellisa Renteria, Majd Hawwar, Croft Jacob, M Ammar Kalas, Marc J. Zuckerman, Jun Zhang

## Abstract

Cerebral cavernous malformations (CCMs) are neurological disorders that make individuals more susceptible to hemorrhagic stroke. The Mexican-Hispanic population has a higher prevalence of both CCMs and metabolic syndrome (MetS), which is also associated with hemorrhagic stroke. A study was conducted with 184 Mexican-Hispanic CCM subjects and age- and sex-matched Hispanic and non-Hispanic white controls. The CCM cohort had a higher proportion of epilepsy and hemorrhagic stroke but a lower proportion of MetS. Higher blood pressure and fasting glucose levels were observed in the CCM cohort. MetS and epilepsy were associated with increased odds of hemorrhagic stroke among elderly CCM patients, and increased systolic blood pressure was significantly linked to increased odds of hemorrhagic stroke in the CCM cohort. To minimize the risk of hemorrhagic stroke, it is important to manage blood pressure and comorbidities like MetS and epilepsy in CCM patients, particularly those older than 50.

**SUMMARUY:** *What is already known about this subject:*

- Metabolic Syndrome (MetS) is widely associated with cardiovascular conditions, including stroke,
- The association between MetS and ischemic stroke is well established
- The relationship between MetS and hemorrhagic stroke remains unclear
- Currently, one two studies explored the relationship between MetS and hemorrhagic stroke in CCM patients; one n sporadic CCM (sCCM) and one in familial CCM cases, with inconsistent results.

*What are the new findings:*

- In this study, a strong association was observed between systolic blood pressure (SBP) and hemorrhagic stroke in the CCM cohort, independent of obesity or fasting glucose levels.
- This study also suggests that MetS is associated with hemorrhagic stroke among CCM patients, specifically in the older age group.

*How might it impact on clinical practice in the foreseeable future:*

- This study demonstrates many of the unique characteristics of symptomatic CCMs within the Mexican-Hispanic population
- Our results suggests that of Mexican-Hispanic CCM subjects are at a greater risk for hemorrhagic stroke and epilepsy than other ethnic groups.
- This study highlights the importance of reducing blood pressure and managing comorbidities such as MetS and epilepsy in CCM patients, especially those who are older than 50 years to minimize the risk of hemorrhagic stroke among CCM subjects

## INTRODUCTION

Cerebral cavernous malformations (CCMs) are neurological disorders characterized by abnormal dilation of intracranial capillaries in the brain, which increases susceptibility to hemorrhagic stroke (1–4). As an inherited condition, familial cases of CCMs (fCCMs) have been linked to three specific genes, CCM1, CCM2, and CCM3 (5–10), while the causation of sporadic cases of CCMs (sCCMs) is still under active investigation (11, 12). Hemorrhagic CCMs are often caused by defects in the blood-brain barrier (BBB), which can result in microvessel rupture (10, 13). The Hispanic population has a higher prevalence and more severe forms of this condition, which is significant because of the greater burden of mortality, disability, and cost associated with hemorrhagic stroke in this population (14–16). Moreover, the Hispanic population is a heterogeneous group and represents the different prevalence of risk factors according to Hispanic origin. Mexican Hispanics, particularly those living on the US-Mexico border have a higher prevalence of metabolic abnormalities and diabetes predisposing them to a greater risk for cardiovascular diseases including stroke compared to Mexican-Hispanic and non-Hispanic white populations. Therefore, it is critical to estimate the prevalence of hemorrhagic stroke in the Mexican-Hispanic CCM cohort compared to Hispanic and non-Hispanic white controls.

Further research is also necessary to fully comprehend the primary risk factors for hemorrhagic stroke in Hispanics, although metabolic syndrome (MetS) and diabetes could be potential contributing factors based on current investigations (14, 15, 17). Evidence indicated that fCCM patients with deficient CCMs proteins, specifically those with *CCM1* gene deficiency, have exhibited disrupted metabolic function, which usually leads to oxidative stress and inflammatory response (18–21). Developmental venous anomalies (DVAs), which are highly associated with sCCMs (22–25), have also been linked to MetS (26–28). However, only a few and inconsistent results have been published regarding the association between MetS and hemorrhagic CCMs (29, 30). Obesity was found to be a significant risk factor for sCCM hemorrhagic events in a large European sCCM cohort (29), while fCCM hemorrhage was only borderline associated with obesity and systolic blood pressure (SBP) in a large Mexican Hispanic population (30). However, it is unclear if MetS or SBP is associated with hemorrhagic stroke among CCM patients or not after adjusting for obesity.

In this study, we sought to estimate the prevalence of hemorrhagic stroke and associated risk factors in the Mexican-Hispanic CCM compared to Hispanic and non-Hispanic white non-CCM controls. We also investigate whether MetS or SBP is independently associated with hemorrhagic stroke in a cohort of Mexican Hispanic subjects with an admixture of sCCM and fCCM cases. By performing a comparative analysis of key components associated with MetS in this population compared to age- and gender-matched healthy individuals, we hope to gain a better understanding of the association between the risk factors and hemorrhagic stroke.

## MATERIALS and METHODS

### Study Population

The El Paso CCM Retrospective Study Part-1 (ElPasoCCMRetro1) database consists of 184 symptomatic CCM subjects identified through a retrospective chart review of patients with ICD-9/10 diagnosis codes covering the CCM clinical spectrum (Suppl. Table 1), familial history, and CT and MRI diagnostics from 2010 to 2023. Data was collected from Texas Tech University Health Sciences Center El Paso clinics and University Medical Center, El Paso (UMC). This study was conducted in accordance with an approved protocol (E22031) from the institutional review board (IRB) at Texas Tech University Health Sciences Center El Paso (TTUHSCEP). We retrospectively obtained variables of interest from patient charts for both familial (fCCMs) and sporadic cases (sCCMs) of CCMs registered at TTUHSCEP/UMC. The physicians’ diagnoses and research staff followed the inclusion criteria for symptomatic CCMs,

### Data acquisition

Data was collected by six trained researchers, mostly medical students supervised by medical residents or postdoctoral fellows. An independent researcher entered the data into the database for standardization and quality control. The database was reviewed for inconsistencies and omissions, and any inconsistent or omitted data points were checked against the original data gathering sheet or the medical record itself. Subjects with inconsistent records were excluded (Suppl. Table 1). If data was unavailable, it was treated as absent, and the subject was considered not to have a condition based on the chart not noting the subject had the condition. Once completed, the database was reviewed again, and subjects were excluded based on criteria such as incomplete medical records, traumatic or infectious stroke etiology, post-operative stroke etiology, presence of brain tumor, no radiographic evidence of infarct, Hispanic birthplace outside the border region (not of Mexican descent), Asian, Black, Native American, or White origin. The remaining subjects in the database were all symptomatic CCM subjects from 2010 to 2023 from the El Paso border region, with a complete medical history and of non-Hispanic White or Mexican-Hispanic ethnicity.

### Outcome

the primary outcome of this study was a hemorrhagic stroke confirmed based on the ICD-9 or 10 codes included in Supplementary Table 1.

### Exposure and risk factors

The primary exposure of interest was MetS which was defined as the presence of three or more of the following: central obesity, elevated triglycerides, reduced high-density lipoprotein (HDL), dyslipidemia, systemic hypertension, or elevated fasting glucose (31). With this classification and well established MetS clinical diagnosis guidelines (32), subjects with MetS were identified in the ElPasoCCMRetro1 database. Some major parameters of metabolic syndrome (MetS), such as SBP, diastolic blood pressure (DBP), blood glucose levels, and body mass index (BMI) were also incuded in this study. In addition to MetS, another critical risk factor was included as epilepsy. Other covariates include age and sex. BMI was calculated using the standard formula in kg/m2.

### Non-CCM controls

For the selection of non-CCM controls, we used a well-established nationally representative non-institutionalized National Health and Nutritional Examination Survey (NHANES) database. We used the most recent and publicly available pre-pandemic cycle 2017-2018 NHANES database for the selection of controls. Initially from this cycle of NHANES, we included all the Mexican-Hispanics (N=1367) and non-Hispanic whites (N= 3150). After removing non-adults, there were 792 Mexican-Hispanics and 2,032 non-Hispanic whites were eligible for control selection. A propensity scores model was developed to identify 2 age- and sex-matched controls per each CCM subject (1:2 case and control selection). Therefore, we selected a total of 368 Mexican Hispanics and 368 non-Hispanic whites. We followed the exact definition of MetS as specified in the above section and defined MetS using the NHANES-based calculations (33, 34). The NHANES collects data from participants on prescribed medications in the past 30 days. We defined epilepsy if participants had taken medications for “epilepsy and recurrent seizures”.

### Data Analysis

Baseline characteristics were summarized using appropriate summary measures including mean and standard deviation (SD) for continuous variables and frequency and percentage for categorical variables. All the characteristics were compared according to the presence and absence of MetS using either Fisher’s exact test or unpaired student’s t-test., The unadjusted and adjusted association of each factor with the presence of hemorrhagic stroke among the CCM cohort was determined using logistic regression analysis. Multiple multivariable logistic regression models were developed, including a model with metabolic syndrome and a model with individual components of metabolic syndrome. In the multivariable model, effect modifiers for the association between MetS and hemorrhagic stroke were explored. In the presence of a strong modifying effect of age, the final logistic regression model was developed for patients with age ≥ 50 years.

Furthermore, the distribution of hemorrhagic stroke and risk factors between the Hispanic CCM cohort and the age- and sex-matched healthy controls were compared using appropriate statistical tests, such as Fisher’s exact test or unpaired student’s t-test. The age- and sex-matched controls were observed using propensity scores logistic regression models, and multivariable logistic regression models were developed to identify differences in risk factors between the CCM cohort and controls. The results of logistic regression models were summarized with the odds ratio (OR), 95% confidence interval (CI), and p-value. P-values less than 5% were considered statistically significant results. The continuous variables in the regression analysis were z-standardized. Statistical analyses were carried out using STATA 17. We followed the conduct and reporting of statistical analysis guidelines in medical research (35, 36).

## RESULTS

### Characteristics of ElPaso CCMRetro1 cohort

A total of 184 CCM cases were included in the study analysis. The average age of the CCM cohort patients was 52.8 (SD: 17.2) years and included 53% females. In the total CCM cohort, 36.6% (n=67) had a hemorrhagic stroke. Approximately 25% (n=45) of patients were identified with epilepsy and 14% (n=25) with MetS. In the CCM cohort, there were no differences in age, sex, or blood pressure between the CCM patients with and without MetS. As obvious, MetS subjects had higher blood glucose concentrations with increased BMI levels (Table 1). Although not statistically significant, there was a trend towards a higher prevalence of hemorrhagic stroke in patients with MetS compared to those without (52 vs. 33.8, p=0.11).

**Table 1.** Distribution of characteristics of Hispanic CCM patients in the entire cohort and by metabolic syndrome (MetS).

| Parameters | Total Cohort | MetS (No) | MetS(Yes) | p-value |
| --- | --- | --- | --- | --- |
| Sample size (N) | 184 | 154 | 25 |  |
| Age (years), mean (SD) | 52.8 (17.2) (n=184) | 51.9 (17.4) | 56.2 (14.9) | 0.24 |
| Sex |  |  |  | 0.67 |
| Male | 86 (46.7%) | 71 (46.1%) | 13 (52.0%) |  |
| Female | 98 (53.3%) | 83 (53.9%) | 12 (48.0%) |  |
| SBP, mean (SD) | 134.3 (23.0) (n=175) | 132.4 (21.6) | 139.6 (26.9) | 0.14 |
| DBP, mean (SD) | 76.0 (13.2) (n=175) | 75.7 (13.7) | 76.2 (10.0) | 0.85 |
| Blood Glucose Levels, mean (SD) | 135.3 (85.4) (n=145) | 126.2 (55.1) | 178.7 (170.3) | 0.007 |
| BMI (kg/m<sup>2</sup>), mean (SD) | 29.1 (6.2) (n=169) | 28.1 (5.2) | 35.0 (8.2) | <0.001 |
| Metabolic syndrome |  |  |  |  |
| No | 154 (83.7%) |  |  |  |
| Yes | 25 (13.6%) |  |  |  |
| Missing | 5 (2.7%) |  |  |  |
| Epilepsy |  |  |  | 0.63 |
| No | 138 (75.0%) | 114 (74.0%) | 20 (80.0%) |  |
| Yes | 45 (24.5%) | 40 (26.0%) | 5 (20.0%) |  |
| Missing | 1 (0.5%) |  |  |  |
| Hemorrhagic Stroke |  |  |  | 0.11 |
| No | 116 (63.0%) | 102 (66.2%) | 12 (48.0%) |  |
| Yes | 67 (36.4%) | 52 (33.8%) | 13 (52.0%) |  |
| Missing | 1 (0.5%) |  |  |  |

BMI; body mass index; SBP: systolic blood pressure; DBP: diastolic blood pressure; SD: standard deviation

### Unadjusted association of MetS and risk factors with hemorrhagic stroke in ElPaso CCMRetro1

For the unadjusted association of covariates with hemorrhagic stroke status among Hispanic CCM patients, only increasing SBP was associated with increased odds of hemorrhagic stroke (OR=1.02, p=0.003). Although not statistically significant, the presence of MetS (OR=2.13) and epilepsy (OR=1.55, p=0.211) also increased the odds of hemorrhagic stroke (Table 2). We observed a strong interaction between age and MetS (OR=1.09, p=0.029) in association with hemorrhagic stroke. MetS increased the odds of hemorrhagic stroke among individuals in the age >50 years group (OR=18.98, p=0.18, Suppl. Table 2). However, the odds of hemorrhagic stroke was not associated with MetS or epilepsy among individuals with two younger age groups (age <35 years or 35-50 years) (Suppl. Table 3). The prevalence of hemorrhagic stroke was significantly higher in MetS patients compared to patients without MetS (70.6% vs. 32.6%) among older patients (Fig. 1). Similarly, hemorrhagic stroke was also found to be higher in the epilepsy group than in the non-epileptic group, especially in older CCM individuals (Fig. 2).

**Figure 1.**
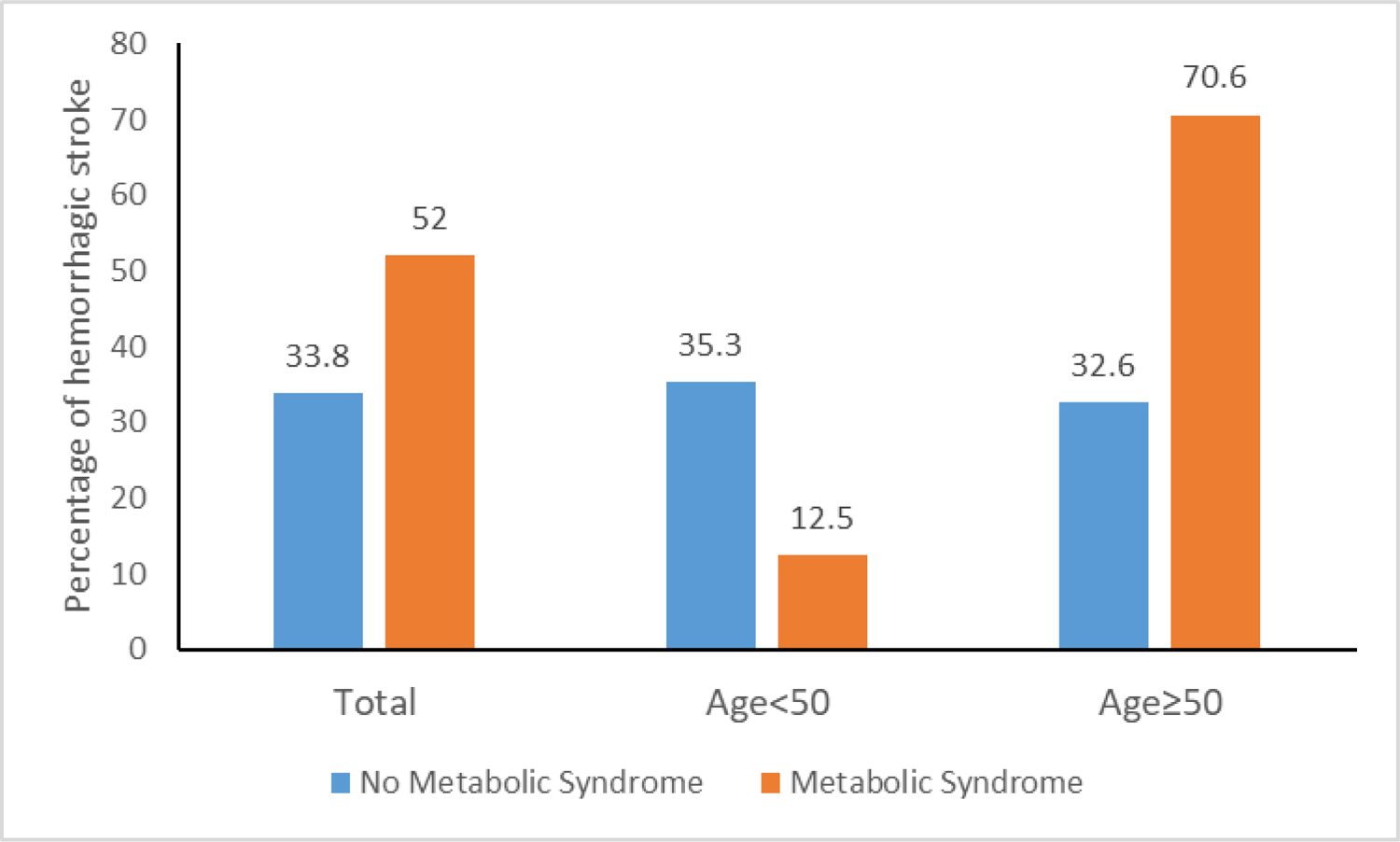
The prevalence of hemorrhagic stroke varies with metabolic syndrome (MetS) status and age in the Hispanic CCM cohort. Comparison of hemorrhagic stroke according to metabolic syndrome status by two age groups in Mexican-Hispanic Cerebral Cavernous Malformations (CCM) cohort.

**Figure 2.**
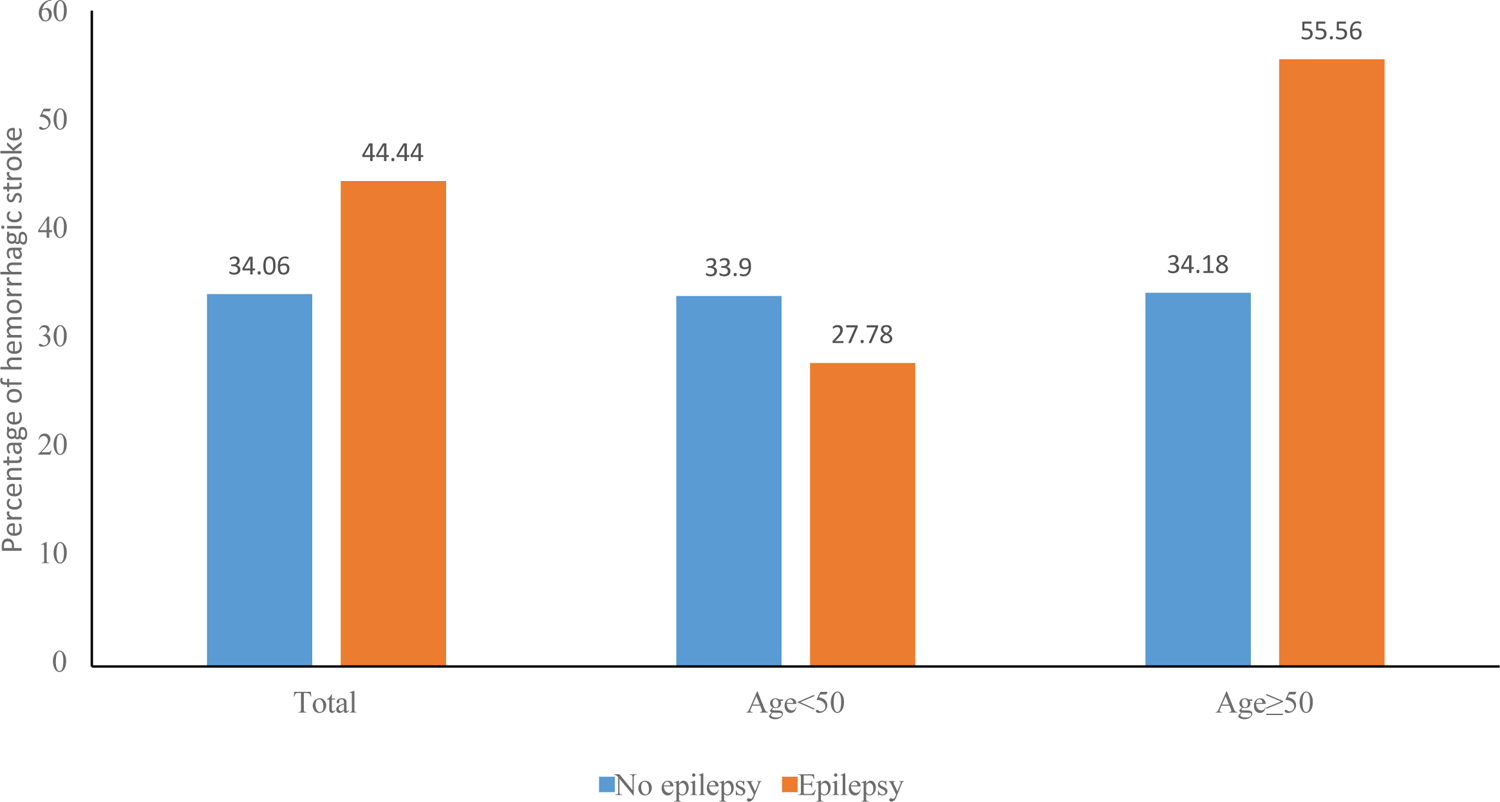
The prevalence of epilepsy varies with metabolic syndrome (MetS) status and age in the Hispanic CCM cohort. Comparison of epilepsy according to metabolic syndrome status by two age groups in Mexican-Hispanic Cerebral Cavernous Malformations (CCM) cohort.

**Table 2.**
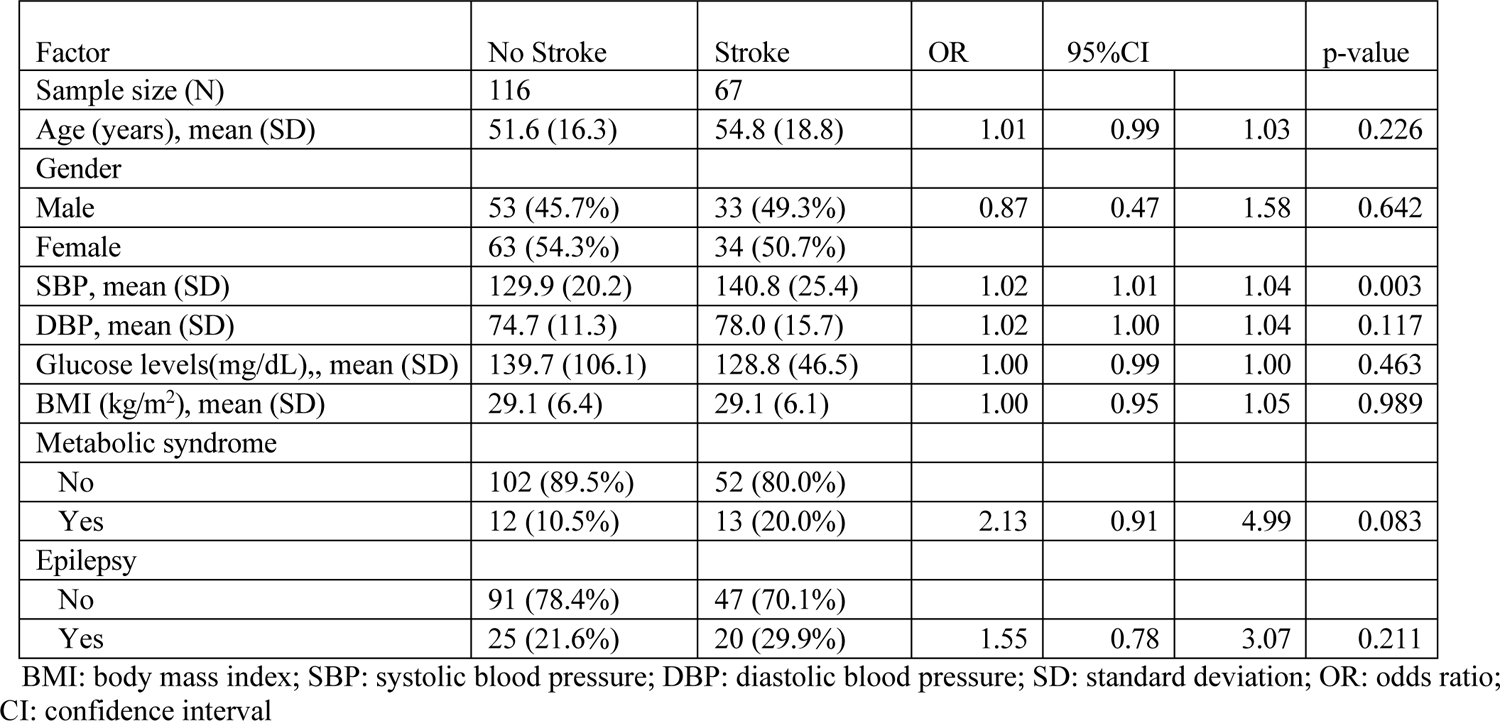
Unadjusted association of covariates with hemorrhagic stroke status among Hispanic CCM patients.

### Unadjusted and adjusted associations of MetS and risk factors with hemorrhagic stroke in older CCM patients

In the unadjusted analysis of CCM patients of old age group (aged 50 years or older), MetS (OR=4.97, p=0.006) and epilepsy (OR=2.41, p=0.05) were found to be significantly associated with hemorrhagic stroke. In the adjusted analysis epilepsy (OR=3.12, p=0.025) and MetS (OR=6.38, p=0.006) remained significant risk factors for hemorrhagic stroke among CCM patients in the older age group (age >=50 years) (Table 3, Fig. 3). In the individual components of MetS analysis, higher SBP remained significantly associated with hemorrhagic stroke (OR=1.54, p=0.025) after adjusting for other cofactors in the model (Fig. 4) and the association remained unchanged after adjusting for DBP instead of fasting glucose levels (Suppl. Table 3).

**Figure 3.**
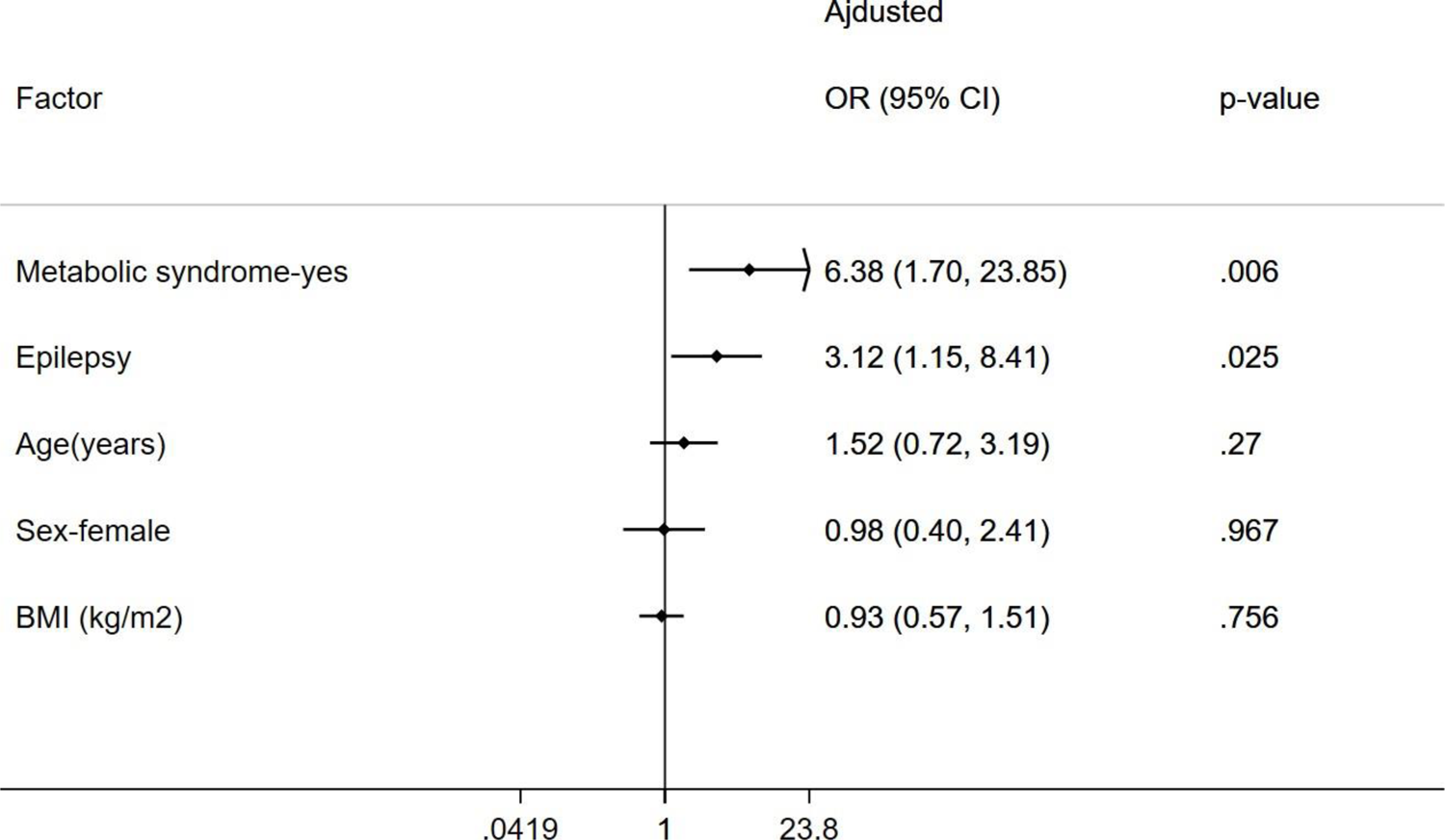
The association between the occurrence of hemorrhagic stroke and the status of MetS and epilepsy in Mexican-Hispanic CCM patients. The significant association between the occurrence of hemorrhagic stroke and the status of MetS and epilepsy in Mexican-Hispanic CCM patients was only found in the older age group (≥50 years), after adjusting for other factors. Z-standardized form of continuous variables included in the regression analysis.

**Figure 4.**
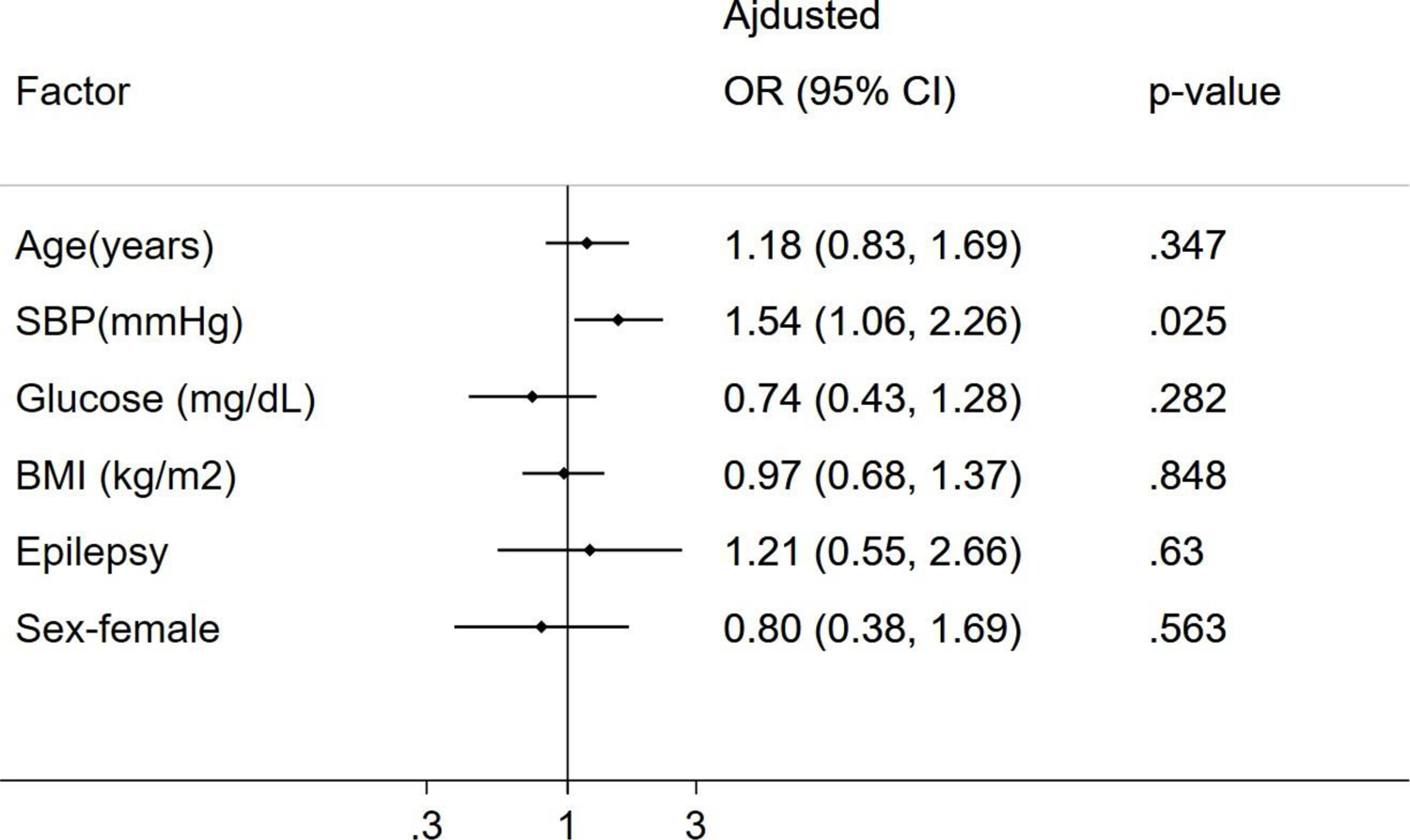
The association between the occurrence of hemorrhagic stroke and blood pressure in Mexican-Hispanic CCM patients. A significant association between the occurrence of hemorrhagic stroke and systolic blood pressure (SBP) in Mexican-Hispanic CCM patients was found, after adjusting for other factors. Z-standardized form of continuous variables included in the regression analysis.

**Table 3.**
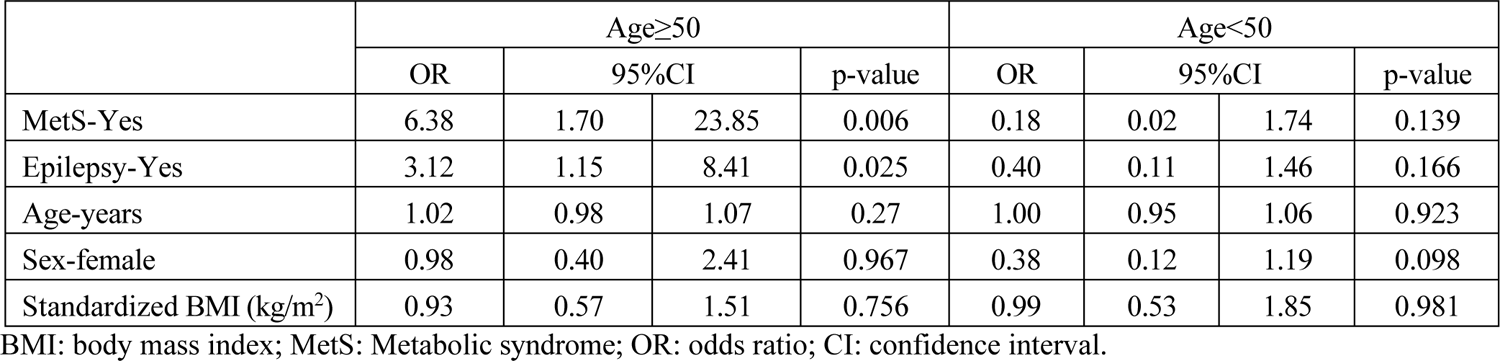
Adjusted association of MS with hemorrhagic stroke status among Hispanic CCM patients of older age group (>=50 years).

|  | Age≥50 |  |  |  | Age<50 |  |  |  |
| --- | --- | --- | --- | --- | --- | --- | --- | --- |
|  | OR | 95%CI |  | p-value | OR | 95%CI |  | p-value |
| MetS-Yes | 6.38 | 1.70 | 23.85 | 0.006 | 0.18 | 0.02 | 1.74 | 0.139 |
| Epilepsy-Yes | 3.12 | 1.15 | 8.41 | 0.025 | 0.40 | 0.11 | 1.46 | 0.166 |
| Age-years | 1.02 | 0.98 | 1.07 | 0.27 | 1.00 | 0.95 | 1.06 | 0.923 |
| Sex-female | 0.98 | 0.40 | 2.41 | 0.967 | 0.38 | 0.12 | 1.19 | 0.098 |
| Standardized BMI (kg/m<sup>2</sup>) | 0.93 | 0.57 | 1.51 | 0.756 | 0.99 | 0.53 | 1.85 | 0.981 |

BMI: body mass index; MetS: Metabolic syndrome; OR: odds ratio; CI: confidence interval.

### Comparison between ElPasoCCMRetro1 CCMs and Mexican Hispanic Controls

In the comparison of age- and gender-matched Mexican Hispanic controls, the Mexican-Hispanic CCM cases had a significantly higher proportion of epilepsy (1.6% vs. 24.6%, p<0.001), hemorrhagic stroke (3.6% vs. 36.6%, p<0.001) but less MetS (54.8% vs. 14%, p<0.001). Among individual components of MetS, Mexican-Hispanic CCM cohort patients had increased blood pressure levels (both SBP and DBP) but decreased BMI levels than age- and gender-matched Hispanic healthy controls (Table 4, Figure 5). Similarly, Mexican-Hispanic CCM cases had a significantly higher proportion of epilepsy (0.8% vs. 24.6%, p<0.001), hemorrhagic stroke (4.8% vs. 36.6%, p<0.001) but less MetS (44.1% vs. 14%, p<0.001) compared to non-Hispanic white controls. Among individual components of MS, Hispanic CCM cohort patients had increased blood pressure levels including fasting glucose levels than non-Hispanic white controls (Table 4, Figure 6).

**Figure 5.**
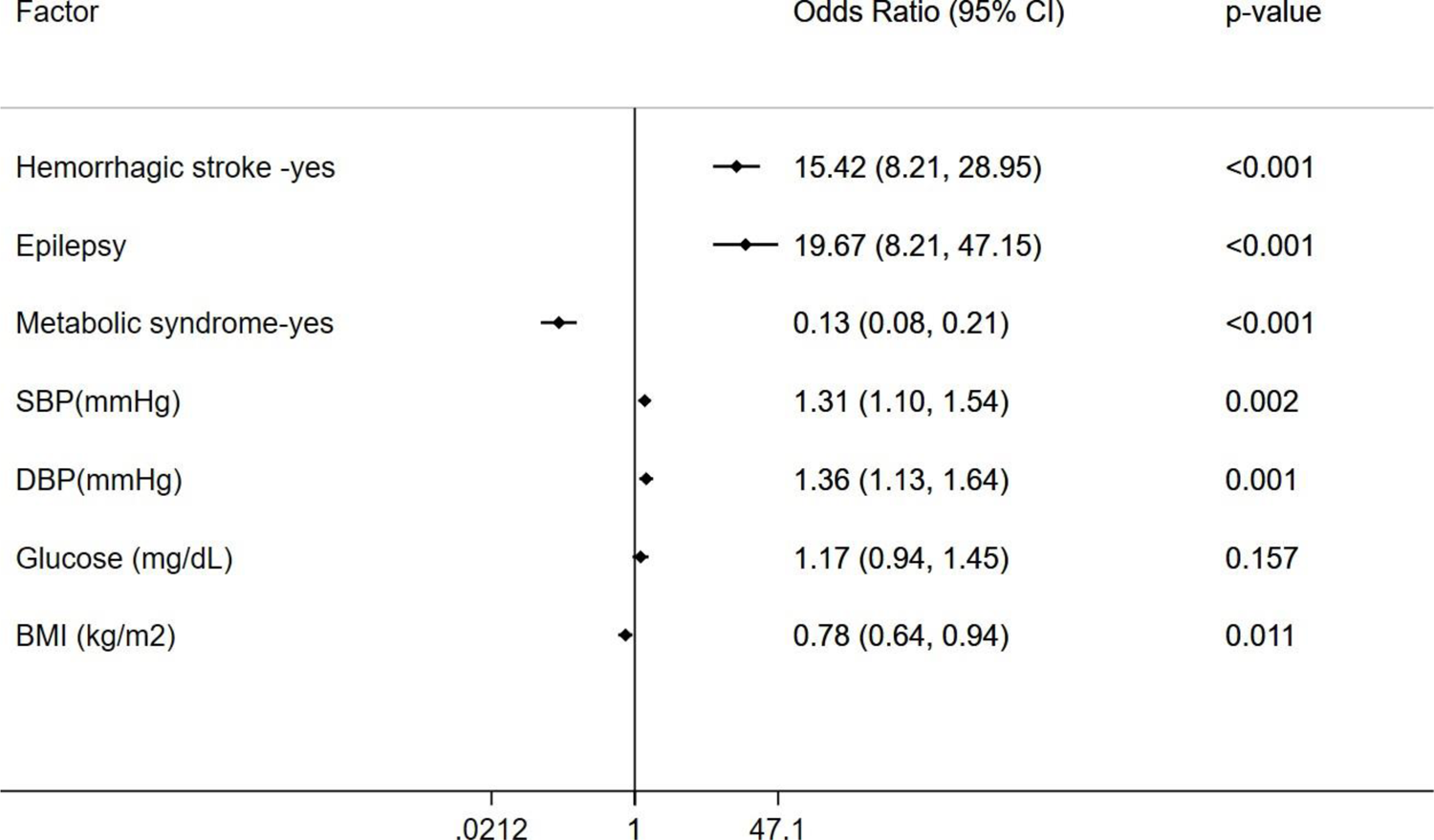
Association between each covariate in the Mexican-Hispanic CCM cohort and age- and gender-matched Mexican-Hispanic controls. The unadjusted correlation between each covariate in the Mexican-Hispanic CCM cohort in comparison to that of the gender- and age-matched Mexican-Hispanic controls from the NHANES dataset (2017-2018) indicate their significant differences.

**Figure 6.**
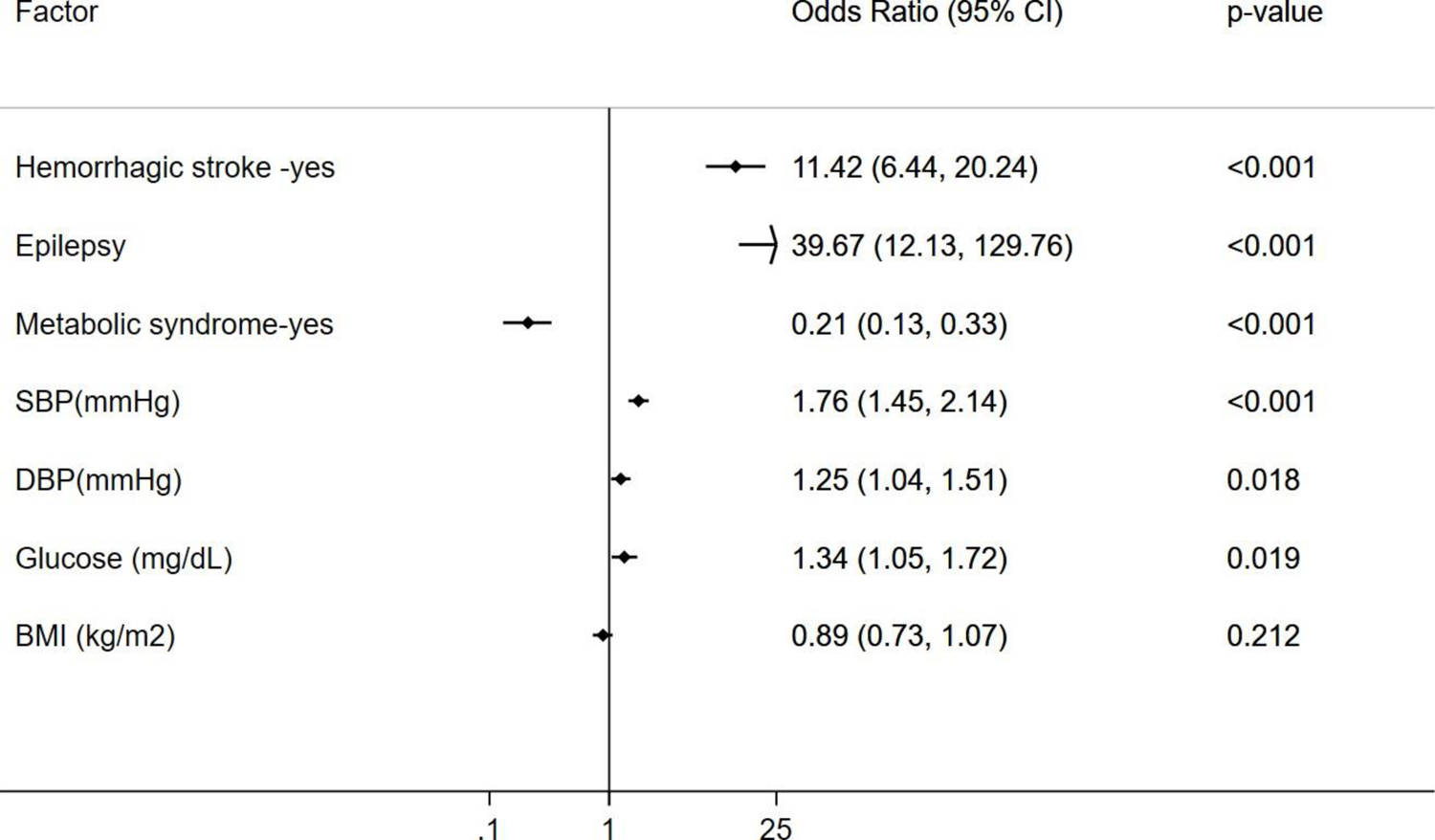
Association between each covariate in the Mexican-Hispanic CCM cohort and age- and gender-matched non-Hispanic white controls. The unadjusted correlation between each covariate in the Mexican-Hispanic CCM cohort in comparison to that of the gender- and age-matched non-Hispanic white controls from the NHANES dataset (2017-2018) indicate their significant differences.

**Table 4.** Comparison of Hispanic CCM cohort with Hispanic controls from NHANES (2017-2018).

| Factor | Mexican-Hispanic CCM | Mexican-Hispanic Controls | Non-Hispanic white Controls | p-value* | p-value** |
| --- | --- | --- | --- | --- | --- |
| N | 184 | 368 | 368 |  |  |
| Age in years, mean (SD) | 52.8 (17.2) | 52.5 (16.5) | 50.7 (16.2) | 0.84 | 0.17 |
| Gender |  |  |  | 1.00 | 0.53 |
| Male | 86 (46.7%) | 171 (46.5%) | 184 (50.0%) |  |  |
| Female | 98 (53.3%) | 197 (53.5%) | 184 (50.0%) |  |  |
| Epilepsy |  |  |  | <0.001 | <0.001 |
| No | 138 (75.4%) | 362 (98.4%) | 365 (99.2%) |  |  |
| Yes | 45 (24.6%) | 6 (1.6%) | 3 (0.8%) |  |  |
| SBP, mean (SD) | 134.3 (23.0) | 127.8 (20.7) | 122.9 (17.3) | 0.002 | <0.001 |
| DBP, mean (SD) | 76.0 (13.2) | 72.2 (11.2) | 73.3 (11.1) | <0.001 | 0.016 |
| Fasting glucose (mg/dL), median (IQR) | 113.0 (97.0, 146.0) | 109.5 (99.0, 128.5) | 102.0 (96.0, 117.0) | 0.46 | 0.005 |
| BMI (kg/m<sup>2</sup>), mean (SD) | 29.1 (6.2) | 30.7 (6.5) | 30.0 (8.2) | 0.011 | 0.21 |
| Metabolic syndrome |  |  |  | <0.001 | <0.001 |
| No | 154 (86.0%) | 163 (45.2%) | 198 (55.9%) |  |  |
| Yes | 25 (14.0%) | 198 (54.8%) | 156 (44.1%) |  |  |
| Stroke |  |  |  | <0.001 | <0.001 |
| No | 116 (63.4%) | 347 (96.4%) | 336 (95.2%) |  |  |
| Yes | 67 (36.6%) | 13 (3.6%) | 17 (4.8%) |  |  |

BMI: body mass index; SBP: systolic blood pressure; DBP: diastolic blood pressure; SD: standard deviation; OR: odds ratio; CI: confidence interval; NHANES: National Health and Nutrition Examination Survey
\*P-values for comparison between Mexican-Hispanic CCM with Mexican-Hispanic controls; \*\* P-values for comparison between Mexican-Hispanic CCM with non-Hispanic white controls

In sum, we found that higher systolic blood pressure (SBP) was significantly associated with hemorrhagic stroke in the CCM cohort even after adjusting for other factors. The CCM cohort had a higher proportion of epilepsy and hemorrhagic stroke but a lower proportion of MetS compared to age- and gender-matched healthy controls. The CCM cohort also had higher levels of blood pressure (both SBP and DBP) and fasting glucose levels, but lower BMI levels compared to both Hispanic and non-Hispanic white controls.

## DISCUSSION

CCMs are neurological disorders characterized by abnormal dilation of intracranial capillaries in the brain, which increases susceptibility to hemorrhagic stroke (1–4). Having an earlier age of onset, a greater incidence, and recurrence of hemorrhagic stroke, Mexican Hispanics have more severe clinical presentations of the CCMs than any other ethnic group (14–16). Therefore, there is an unmet need to better understand the characteristics of risk factors for hemorrhagic stroke in the Mexican-Hispanic population. Our study found that Mexican-Hispanic CCMs had a higher prevalence of hemorrhagic stroke and epilepsy but a lower proportion of MetS compared to age- and sex-matched Mexican-Hispanic and non-Hispanic white controls. Elevated blood pressure and glucose levels but not higher BMI were the primary risk factors in the CCM cohort compared to non-CCM controls and higher glucose levels were identified as the independent risk factor for hemorrhagic stroke in CCM patients. In our study, the presence of MetS and epilepsy was associated with hemorrhagic stroke among CCM patients who are older than 50 years.

It is widely known that MetS is associated with cardiovascular conditions (31, 37–39) and there has been a long-standing exploration of the potential link between MetS and stroke (37, 38, 40–42). Various study designs have been used to investigate this association (43–45), but most studies have focused on the most common form of stroke, ischemic stroke, in various ethnic groups (38, 39, 46–49), including the Mexican Hispanic population (50, 51). Although the association between MetS and ischemic stroke has been well established, the relationship between MetS and hemorrhagic stroke remains unclear (44, 48). Furthermore, metabolic dysfunction and its resulting consequences, such as oxidative stress, and inflammatory responses, have been extensively documented in conditions with CCMs deficiency (18–21), particularly in cases of CCM1 gene deficiency in fCCMs (18, 21, 52). This suggests a potential link between metabolic dysfunction and CCMs, but more research is needed to fully understand this relationship. Our study suggests that MetS is associated with hemorrhagic stroke among CCM patients but in the older age group. Notably, developmental venous anomalies (DVA), the most frequent type of neurovascular malformation linked to the pathogenesis of sporadic CCMs (22–25), have been identified as being associated with MetS (26–28). A five-year prospective study has identified DVA as one of the most significant risk factors for hemorrhagic events in brainstem CCMs (53), indirectly confirming our study findings that MetS is associated with sCCM hemorrhage. However, a 15-year retrospective study of a large cohort of 682 sCCM patients found that obesity may be the only risk factor for sCCM hemorrhagic events, as no significant association was found between sCCM hemorrhage and other key components of MetS, such as hypertension, diabetes, and hyperlipidemia (29). The differences may be observed due to unexplored interaction between age and MetS on hemorrhagic stroke in previous CCM cohort analyses. A three-year cross-sectional study involving a large cohort of 185 fCCM patients with the common Hispanic CCM1 mutation found no significant association between CCM lesion count and diabetes or hyperlipidemia, but borderline associations of fCCM hemorrhage with obesity and SBP were observed (30). We also observed a strong association between SBP and hemorrhagic stroke in our CCM cohort independent of obesity or fasting glucose levels.

Although epilepsy has been a known factor associated with hemorrhagic stroke regardless of the ethnic population, our study also observed a strong association between epilepsy and hemorrhagic stroke in older CCM subjects only. Multiple studies confirm that epilepsy is profoundly associated with hemorrhagic stroke in older adults (54). Furthermore, the CCM cohort demonstrated an increased prevalence of epilepsy compared to controls confirming that CCM subjects are at risk for epilepsy and related diseases. In fact, post-stroke seizures and epilepsy is highly prevalent in older adults that subsequently induce metabolic stress and mortality (55). Our CCM cohort had a much lower prevalence of MetS compared to non-CCM Hispanic or non-Hispanic controls, despite Hispanics having the highest prevalence of MetS than any other ethnic population. This suggests that symptomatic CCMs increase the risk of stroke by increasing hypertension and glucose rather than obesity as identified in our study. High glucose levels and hypertension increase oxidative stress and uric acid leading to reduce vascular relaxation through inhibiting nitric oxide and elevating the risk of hemorrhagic stroke (56–58).

One of the major limitations of our study is the cross-sectional study design. Owing to the non-follow-up study, we cannot establish a causal link between hemorrhagic strokes and MetS. Furthermore, due to the retrospective nature of the study, we could not collect several risk factors associated with metabolic health and stroke in our study. Regardless of this limitation, our study is the first study characterizing hemorrhagic stroke, MetS, and epilepsy among a homogenous cohort of Hispanic CCM subjects. The inclusion of two age- and sex-matched ethnic controls is another strength of our study allowing us to identify the differences in CCM characteristics compared to non-CCM subjects.

Our results demonstrate many of the unique characteristics of symptomatic CCMs within the Mexican-Hispanic population of the El Paso border region. In our study, more than one-third of Mexican-Hispanic CCM subjects had a hemorrhagic stroke. In addition, one-fourth of the Mexican-Hispanic CCM subjects had epilepsy. Our study clearly suggests that of Mexican-Hispanic CCM subjects are at a greater risk for hemorrhagic stroke and epilepsy than other ethnic controls. Increasing systolic blood pressure is an independent factor associated with hemorrhagic stroke in the CCM cohort. Among CCM subjects of age greater than or equal to 50 years, metabolic syndrome and epilepsy were identified as the risk factors for hemorrhagic stroke. Overall, the study highlights the importance of reducing blood pressure and managing comorbidities such as MetS and epilepsy in CCM patients, especially those who are older than 50 years to minimize the risk of hemorrhagic stroke among CCM subjects.

## Data Availability

All data produced in the present study are available upon reasonable request to the authors.

## Acknowledgements

We would like to thank Johnathan Abou-Fadel, Muaz Bhalli, Revathi Gnanasekaran, Alexander Le, Nickolas Sanchez, Charlie Harvey and Drexell Vincent for their efforts and help with this project.

## Author Contributions

Alok Dwivedi: Methodology, Writing-Original draft preparation, Writing-Reviewing and Editing; David Jang: Experiments-data collection, data curation; Ofek Belkin: Experiments-data collection, data curation and management; Justin Aickareth: Experiments-data collection, data curation; Mellisa Renteria: Experiments-data collection, data curation and management; Majd Hawwar: Experiments-data collection, data curation; Croft, Jacob: Experiments-data curation and management; Writing-Original draft preparation and Illustrating, Ammar M Kalas: Experiments-data collection, data curation, Marc Zuckerman: Methodology, Experiments-data collection, data curation, Jun Zhang: Conceptualization, Methodology, Writing-Original draft preparation, Writing-Reviewing and Editing.

## Online Supplement

Supplementary information is available at the *J Investig Med* website

## References

1. Otten P, Pizzolato GP, Rilliet B, et al. [131 cases of cavernous angioma (cavernomas) of the CNS, discovered by retrospective analysis of 24,535 autopsies]. Neurochirurgie. 1989; 35:82–83, 128-131.

2. Moriarity JL, Wetzel M, Clatterbuck RE, et al. The natural history of cavernous malformations: a prospective study of 68 patients. Neurosurgery. 1999; 44:1166–1171; discussion 1172-1163.

3. Del Curling O, Jr., Kelly DL, Jr., Elster AD, et al. An analysis of the natural history of cavernous angiomas. J Neurosurg. 1991; 75:702–708.

4. Batra S, Lin D, Recinos PF, et al. Cavernous malformations: natural history, diagnosis and treatment. Nat Rev Neurol. 2009; 5:659–670.

5. Rigamonti D, Hadley MN, Drayer BP, et al. Cerebral cavernous malformations. Incidence and familial occurrence. N Engl J Med. 1988; 319:343–347.

6. Rigamonti D, Drayer BP, Johnson PC, et al. The MRI appearance of cavernous malformations (angiomas). J Neurosurg. 1987; 67:518–524.

7. Johnson EW, Iyer LM, Rich SS, et al. Refined localization of the cerebral cavernous malformation gene (CCM1) to a 4-cM interval of chromosome 7q contained in a well-defined YAC contig. Genome Res. 1995; 5:368–380.

8. Craig HD, Gunel M, Cepeda O, et al. Multilocus linkage identifies two new loci for a mendelian form of stroke, cerebral cavernous malformation, at 7p15-13 and 3q25.2-27. Hum Mol Genet. 1998; 7:1851–1858.

9. Zhang J, Clatterbuck RE, Rigamonti D, et al. Cloning of the murine Krit1 cDNA reveals novel mammalian 5’ coding exons. Genomics. 2000; 70:392–395.

10. Padarti A and Zhang J. Recent advances in cerebral cavernous malformation research. Vessel Plus. 2018; 2.

11. Zhang J, Abou-Fadel J, Renteria M, et al. Cerebral cavernous malformations do not fall in the spectrum of PIK3CA-related overgrowth. J Neurol Neurosurg Psychiatry. 2022.

12. Peyre M, Miyagishima D, Bielle F, et al. Somatic PIK3CA Mutations in Sporadic Cerebral Cavernous Malformations. N Engl J Med. 2021; 385:996–1004.

13. Clatterbuck RE, Eberhart CG, Crain BJ, et al. Ultrastructural and immunocytochemical evidence that an incompetent blood-brain barrier is related to the pathophysiology of cavernous malformations. J Neurol Neurosurg Psychiatry. 2001; 71:188–192.

14. Rodriguez CJ, Allison M, Daviglus ML, et al. Status of cardiovascular disease and stroke in Hispanics/Latinos in the United States: a science advisory from the American Heart Association. Circulation. 2014; 130:593–625.

15. Cruz-Flores S, Rabinstein A, Biller J, et al. Racial-ethnic disparities in stroke care: the American experience: a statement for healthcare professionals from the American Heart Association/American Stroke Association. Stroke. 2011; 42:2091–2116.

16. Trimble B and Morgenstern LB. Stroke in minorities. Neurol Clin. 2008; 26:1177–1190, xi.

17. Hertz RP, Unger AN and Ferrario CM. Diabetes, hypertension, and dyslipidemia in Mexican Americans and non-Hispanic whites. Am J Prev Med. 2006; 30:103–110.

18. Mastrocola R, Aimaretti E, Ferreira Alves G, et al. Heterozygous Loss of KRIT1 in Mice Affects Metabolic Functions of the Liver, Promoting Hepatic Oxidative and Glycative Stress. Int J Mol Sci. 2022; 23.

19. Retta SF and Glading AJ. Oxidative stress and inflammation in cerebral cavernous malformation disease pathogenesis: Two sides of the same coin. Int J Biochem Cell Biol. 2016; 81:254–270.

20. Antognelli C, Perrelli A, Armeni T, et al. Dicarbonyl Stress and S-Glutathionylation in Cerebrovascular Diseases: A Focus on Cerebral Cavernous Malformations. Antioxidants (Basel). 2020; 9.

21. Choquet H, Trapani E, Goitre L, et al. Cytochrome P450 and matrix metalloproteinase genetic modifiers of disease severity in Cerebral Cavernous Malformation type 1. Free Radic Biol Med. 2016; 92:100–109.

22. Mooney MA and Zabramski JM. Developmental venous anomalies. Handb Clin Neurol. 2017; 143:279–282.

23. Petersen TA, Morrison LA, Schrader RM, et al. Familial versus sporadic cavernous malformations: differences in developmental venous anomaly association and lesion phenotype. AJNR Am J Neuroradiol. 2010; 31:377–382.

24. Idiculla PS, Gurala D, Philipose J, et al. Cerebral Cavernous Malformations, Developmental Venous Anomaly, and Its Coexistence: A Review. Eur Neurol. 2020; 83:360–368.

25. Brinjikji W, El-Masri AE, Wald JT, et al. Prevalence of cerebral cavernous malformations associated with developmental venous anomalies increases with age. Childs Nerv Syst. 2017; 33:1539–1543.

26. Timerman D, Thum JA and Larvie M. Quantitative Analysis of Metabolic Abnormality Associated with Brain Developmental Venous Anomalies. Cureus. 2016; 8:e799.

27. Larvie M, Timerman D and Thum JA. Brain metabolic abnormalities associated with developmental venous anomalies. AJNR Am J Neuroradiol. 2015; 36:475–480.

28. Lazor JW, Schmitt JE, Loevner LA, et al. Metabolic Changes of Brain Developmental Venous Anomalies on (18)F-FDG-PET. Acad Radiol. 2019; 26:443–449.

29. Chen B, Saban D, Rauscher S, et al. Modifiable Cardiovascular Risk Factors in Patients With Sporadic Cerebral Cavernous Malformations: Obesity Matters. Stroke. 2021; 52:1259–1264.

30. Choquet H, Nelson J, Pawlikowska L, et al. Association of cardiovascular risk factors with disease severity in cerebral cavernous malformation type 1 subjects with the common Hispanic mutation. Cerebrovasc Dis. 2014; 37:57–63.

31. Mottillo S, Filion KB, Genest J, et al. The metabolic syndrome and cardiovascular risk a systematic review and meta-analysis. J Am Coll Cardiol. 2010; 56:1113–1132.

32. Alberti KG, Eckel RH, Grundy SM, et al. Harmonizing the metabolic syndrome: a joint interim statement of the International Diabetes Federation Task Force on Epidemiology and Prevention; National Heart, Lung, and Blood Institute; American Heart Association; World Heart Federation; International Atherosclerosis Society; and International Association for the Study of Obesity. Circulation. 2009; 120:1640–1645.

33. Rajkumar P, Dwivedi AK, Dodoo CA, et al. The association between metabolic syndrome and Hepatitis C virus infection in the United States. Cancer Causes Control. 2020; 31:569–581.

34. Dubey P, Reddy SY, Singh V, et al. Association of Exposure to Phthalate Metabolites With Sex Hormones, Obesity, and Metabolic Syndrome in US Women. JAMA Netw Open. 2022; 5:e2233088.

35. Dwivedi AK and Shukla R. Evidence-based statistical analysis and methods in biomedical research (SAMBR) checklists according to design features. Cancer Rep (Hoboken). 2020; 3:e1211.

36. Dwivedi AK. How to write statistical analysis section in medical research. J Investig Med. 2022; 70:1759–1770.

37. Hajhosseiny R, Matthews GK and Lip GY. Metabolic syndrome, atrial fibrillation, and stroke: Tackling an emerging epidemic. Heart Rhythm. 2015; 12:2332–2343.

38. Liang Y, Yan Z, Hao Y, et al. Metabolic syndrome in patients with first-ever ischemic stroke: prevalence and association with coronary heart disease. Sci Rep. 2022; 12:13042.

39. Chen YC, Sun CA, Yang T, et al. Impact of metabolic syndrome components on incident stroke subtypes: a Chinese cohort study. J Hum Hypertens. 2014; 28:689–693.

40. Towfighi A and Ovbiagele B. Metabolic syndrome and stroke. Curr Diab Rep. 2008; 8:37–41.

41. Tang XN, Liebeskind DS and Towfighi A. The Role of Diabetes, Obesity, and Metabolic Syndrome in Stroke. Semin Neurol. 2017; 37:267–273.

42. Rothwell PM. Prevention of stroke in patients with diabetes mellitus and the metabolic syndrome. Cerebrovasc Dis. 2005; 20 Suppl 1:24–34.

43. Li X, Li X, Fang F, et al. Is Metabolic Syndrome Associated with the Risk of Recurrent Stroke: A Meta-Analysis of Cohort Studies. J Stroke Cerebrovasc Dis. 2017; 26:2700–2705.

44. Takahashi T, Harada M, Kikuno T, et al. Prevalence of metabolic syndrome in stroke patients: a prospective multicenter study in Japan. Acute Med Surg. 2014; 1:17–22.

45. Zhang F, Liu L, Zhang C, et al. Association of Metabolic Syndrome and Its Components With Risk of Stroke Recurrence and Mortality: A Meta-analysis. Neurology. 2021; 97:e695–e705.

46. Sarrafzadegan N, Gharipour M, Sadeghi M, et al. Metabolic Syndrome and the Risk of Ischemic Stroke. J Stroke Cerebrovasc Dis. 2017; 26:286–294.

47. Lucke-Wold BP, Turner RC, Lucke-Wold AN, et al. Age and the metabolic syndrome as risk factors for ischemic stroke: improving preclinical models of ischemic stroke. Yale J Biol Med. 2012; 85:523–539.

48. Noda H, Iso H, Saito I, et al. The impact of the metabolic syndrome and its components on the incidence of ischemic heart disease and stroke: the Japan public health center-based study. Hypertens Res. 2009; 32:289–298.

49. Chei CL, Yamagishi K, Tanigawa T, et al. Metabolic Syndrome and the Risk of Ischemic Heart Disease and Stroke among Middle-Aged Japanese. Hypertens Res. 2008; 31:1887–1894.

50. Osborn MF, Miller CC, Badr A, et al. Metabolic syndrome associated with ischemic stroke among the Mexican Hispanic population in the El Paso/US-Mexico border region. J Stroke Cerebrovasc Dis. 2014; 23:1477–1484.

51. Senior D, Osborn MF, Tajnert K, et al. Moderate and Severe Blood Pressure Elevation Associated with Stroke in the Mexican Hispanic Population. Health (Irvine Calif). 2017; 9:951–963.

52. Perrelli A and Retta SF. Polymorphisms in genes related to oxidative stress and inflammation: Emerging links with the pathogenesis and severity of Cerebral Cavernous Malformation disease. Free Radic Biol Med. 2021; 172:403–417.

53. Kong L, Ma XJ, Xu XY, et al. Five-year symptomatic hemorrhage risk of untreated brainstem cavernous malformations in a prospective cohort. Neurosurg Rev. 2022; 45:2961–2973.

54. Galovic M, Ferreira-Atuesta C, Abraira L, et al. Seizures and Epilepsy After Stroke: Epidemiology, Biomarkers and Management. Drugs Aging. 2021; 38:285–299.

55. Mohamed AT, El Rakawy MH, Abdelhamid YAE, et al. Stroke-related early seizures: clinical and neurophysiological study in a sample of Egyptian population. Egypt J Neurol Psych. 2023; 59.

56. Unnithan AKA, J MD and Mehta P. Hemorrhagic Stroke. StatPearls. Treasure Island (FL); 2023.

57. Lee M, Saver JL, Hong KS, et al. Effect of pre-diabetes on future risk of stroke: meta-analysis. Bmj-Brit Med J. 2012; 344.

58. Hu G, Sarti C, Jousilahti P, et al. The impact of history of hypertension and type 2 diabetes at baseline on the incidence of stroke and stroke mortality. Stroke. 2005; 36:2538–2543.

