## Supplementary material for "Metabolic syndrome and hemorrhagic stroke among symptomatic CCMs in the Mexican Hispanic Population": Suppl Materials

### Supplemental Materials

[*]All correspondence:

Jun Zhang, Sc.D., Ph.D.

Molecular and translational Medicine

Texas Tech University Health Sciences Center El Paso

5001 El Paso Dr., El Paso, TX 79905

**Legends**

**Suppl Table 1. Variables of interest for data collection of CCM patients.** All clinical data from the patients admitted to TTUHSCEP/UMC presenting with confirmed or assumed cerebral cavernous malformations (CCMs) will be collected based on variables of interest.

**Suppl. Table 2. An interaction between age and metabolic syndrome (MetS) on hemorrhagic stroke among Mexican -Hispanic CCM patients**

**Suppl. Table 3. Distribution of MetS, epilepsy, and hemorrhagic stroke by age groups among Hispanic CCM patients**

**Suppl Table 1. Variables of interest for data collection of CCM patients.** All clinical data from the patients admitted to TTUHSCEP/UMC presenting with confirmed or assumed cerebral cavernous malformations (CCMs) and MetS parameters will be collected based on variables of interest listed below. (Note: not all risk factors were used in this study).

| |
|---|
| **Age years (EX: 32)** |
| **Date of first-ever ICH (if occurred) (year)** |
| **Date of the first-ever seizure (if occurred) (years)** |
| **Time point of any treatment (neurosurgery, radiosurgery) (if occurred) (year)** |
| **Confirmed familial disease (MRI pattern plus one affected relative or pos. genotyping) (yes/no)** |
| **Assumed familial disease (MRI pattern without an affected relative/genotyping) (yes/no)** |
| **Time point of the last follow-up at the hospital/institution (Year)** |
| **Confirmed mutation (CCM1/2/3) (if available)** |
| **Functional state (mRS) at last FU (if available)** |
| **Stroke Type (Ischemic 1 Hemorrhagic 2)** |
| **Place of Birth (Mexico, El Paso, UNK)** |
| **Admit Date (Year only)** |
| **Hypertension (1, yes; 2, no)** |
| **Hypertension numbers** |
| **Systolic BP at presentation** |
| **Diastolic BP at presentation** |
| **Mean Arterial Pressure number** |
| **Tobacco abuse (1, yes; 2, no)** |
| **Tobacco Info (packs/day)** |
| **Diabetes(1, yes; 2, no)** |
| **Diabetes Info (I/II; glucose Value)** |
| **Heart Disease (1, yes; 2, no)** |

Suppl. Table 2

| Parameters | OR | 95%CI | | p-value |
|---|---|---|---|---|
| MetS | 0.01 | 0.00 | 1.43 | 0.071 |
| Age-years | 1.00 | 0.98 | 1.02 | 0.904 |
| MetS and age | 1.09 | 1.01 | 1.18 | 0.029 |
| MeTS | 0.26 | 0.03 | 2.26 | 0.223 |
| Age≥50 | 0.89 | 0.45 | 1.73 | 0.722 |
| MetS and Age≥50 | 18.98 | 1.66 | 216.70 | 0.018 |

MetS: metabolic syndrome; OR: odds ratio; CI: confidence interval

Suppl. Table 3

| Parameters | Age ≤35 | Age (35-50) | Age≥50 | p-value |
|---|---|---|---|---|
| Sample size (N) | 33 | 44 | 107 | |
| Metabolic syndrome | | | | 0.38 |
| No | 31 (93.9%) | 37 (86.0%) | 86 (83.5%) | |
| Yes | 2 (6.1%) | 6 (14.0%) | 17 (16.5%) | |
| Epilepsy | | | | 0.77 |
| No | 24 (72.7%) | 35 (79.5%) | 79 (74.5%) | |
| Yes | 9 (27.3%) | 9 (20.5%) | 27 (25.5%) | |
| Stroke | | | | 0.60 |
| No | 23 (69.7%) | 29 (65.9%) | 64 (60.4%) | |
| Yes | 10 (30.3%) | 15 (34.1%) | 42 (39.6%) | |
| Hemorrhagic Stroke | OR, p-value | OR, p-value | OR, p-value | |
| MetS-Yes | 0.41, 0.575 | 0.44, 0.405 | 4.67, 0.006 | |
| Epilepsy-Yes | 0.65, 0.606 | 1.01,0.987 | 2.37,0.054 | |

OR: odds ratio.
